# Analysis of the risk and pre-emptive control of viral outbreaks accounting for within-host dynamics: SARS-CoV-2 antigen testing as a case study

**DOI:** 10.1101/2023.03.23.23287633

**Authors:** William S Hart, Hyeongki Park, Yong Dam Jeong, Kwang Su Kim, Raiki Yoshimura, Robin N Thompson, Shingo Iwami

**Author notes:** These authors contributed equally to this work (Robin N Thompson, Shingo Iwami).

## Abstract

In the era of living with COVID-19, the risk of localised SARS-CoV-2 outbreaks remains. Here, we develop a multi-scale modelling framework for estimating the local outbreak risk for a viral disease (the probability that a major outbreak results from a single case introduced into the population), accounting for within-host viral dynamics. Compared to population-level models previously used to estimate outbreak risks, our approach enables more detailed analysis of how the risk can be mitigated through pre-emptive interventions such as antigen testing. Considering SARS-CoV-2 as a case study, we quantify the within-host dynamics using data from individuals with omicron variant infections. We demonstrate that regular antigen testing reduces, but may not eliminate, the outbreak risk, depending on characteristics of local transmission. In our baseline analysis, daily antigen testing reduces the outbreak risk by 45% compared to a scenario without antigen testing. Additionally, we show that accounting for heterogeneity in within-host dynamics between individuals affects outbreak risk estimates and assessments of the impact of antigen testing. Our results therefore highlight important factors to consider when using multi-scale models to design pre-emptive interventions against SARS-CoV-2 and other viruses.

## Introduction

Following the widespread rollout of COVID-19 vaccines, countries worldwide have adopted policies of “living with COVID-19” (for example, the UK removed its final domestic restrictions in February 2022 (1)). Waves of COVID-19 cases continue to occur (2), generated by factors including waning immunity (3,4) and the continued evolution of the SARS-CoV-2 virus (5–7), although vaccines provide high levels of ongoing protection against severe disease. Nonetheless, localised outbreaks, either in geographical areas or in specific populations such as schools, universities and workplaces, continue to cause disruption (for example, through student or staff absence).

Mathematical modelling can be used to estimate the (local) *outbreak risk*, which is defined as the probability that a major infectious disease outbreak results from a single infection occurring within the population (8–12). While the outbreak risk can be estimated by simulating a stochastic epidemic model a large number of times (and calculating the proportion of simulations in which a large outbreak occurs), branching process theory can also be used to derive outbreak risk estimates analytically (12). A commonly used formula in the applied epidemic modelling literature (13–19) is Outbreak risk = 1 − 1/*R*_0_ (whenever the basic reproduction number, *R*_0_ > 1; when *R*_0_ ≤ 1, the outbreak risk is zero). However, this formula relies on simplistic assumptions, including each infected individual having constant infectiousness throughout an exponentially distributed infectious period. Several studies have therefore relaxed these assumptions, for example by considering a gamma-distributed infectious period (14,17,20) and/or accounting for heterogeneity between age groups (9,21,22).

In multi-scale epidemic modelling frameworks, within-host viral dynamics models, which describe how the viral load of an infected host evolves over the course of infection and can be calibrated to longitudinal individual data, are used to inform population-level epidemiological models (23–25). One advantage of such approaches is that they facilitate a detailed description of the impact of interventions, such as antigen testing (26–28) or the use of antiviral drugs (29), which depend upon and/or affect the within-host dynamics in a manner that cannot be fully captured in simple population-level models. Multi-scale approaches have been applied to SARS-CoV-2 (26–33) and other pathogens including influenza (23,34) to generate outbreak projections and test control interventions. However, multi-scale methods have not previously been used to estimate the outbreak risk, or to analyse how the risk can be mitigated through pre-emptive interventions.

In this study, we develop a multi-scale approach for calculating the outbreak risk, accounting for within-host viral dynamics and heterogeneity in these dynamics between individuals. We derive an equation satisfied by the outbreak risk under a multi-scale model, and verify our analytically derived outbreak risk estimates using simulations of an individual-based stochastic outbreak simulation model. Focussing on the case study of SARS-CoV-2, we characterise the viral dynamics by fitting a within-host model (35–42) to data from 521 individuals with infections due to the omicron variant (43). We first consider the outbreak risk in the absence of interventions, before exploring the extent to which the outbreak risk can be mitigated through regular rapid antigen testing of the entire local population. Additionally, we analyse the impact of the reproduction number for local transmissions, the level of transmission following detection, heterogeneity in within-host dynamics, and asymptomatic infection, on the outbreak risk and the effectiveness of antigen testing.

Our results highlight that the impact of regular antigen testing on the local SARS-CoV-2 outbreak risk is dependent on the regularity of testing, as well as the exact population under consideration (including the level of vaccine- or infection-acquired immunity) and the characteristics of the viral variant responsible for infections. Based on our analysis, we expect antigen testing to reduce the outbreak risk but not eliminate it completely. We stress that while SARS-CoV-2 is our focus here, our general approach can be applied to other viruses in preparedness for future outbreaks, epidemics and pandemics beyond COVID-19.

## Results

Our multi-scale modelling framework for estimating outbreak risks and analysing the impact of pre-emptive control is outlined in the context of SARS-CoV-2 and regular antigen testing in **Figure 1**. In our approach, a within-host model is first fitted to individual infection data to estimate the viral load of infected hosts at each time since infection, potentially accounting for heterogeneity in within-host dynamics between different individuals (**Figure 1A**). Accounting for a reduced transmission risk following detection, which may occur prior to symptom onset if regular antigen testing is carried out (**Figure 1B**), the viral load profile(s) can be used to estimate the infectiousness profile(s) (**Figure 1C**). The outbreak risk, following a single newly infected individual arriving in an otherwise uninfected population, is then estimated under a branching process transmission model incorporating the estimated infectiousness profile(s) (**Figure 1D**). Specifically, we have analytically derived equations satisfied by the outbreak risk assuming either homogeneous (Eq. (1) in **Methods**) or heterogeneous (Eq. (2)) within-host dynamics (the derivations are described in **Supplementary Note 1**). The effect on the outbreak risk of factors such as the frequency of antigen testing can then be analysed.

**Figure 1.**
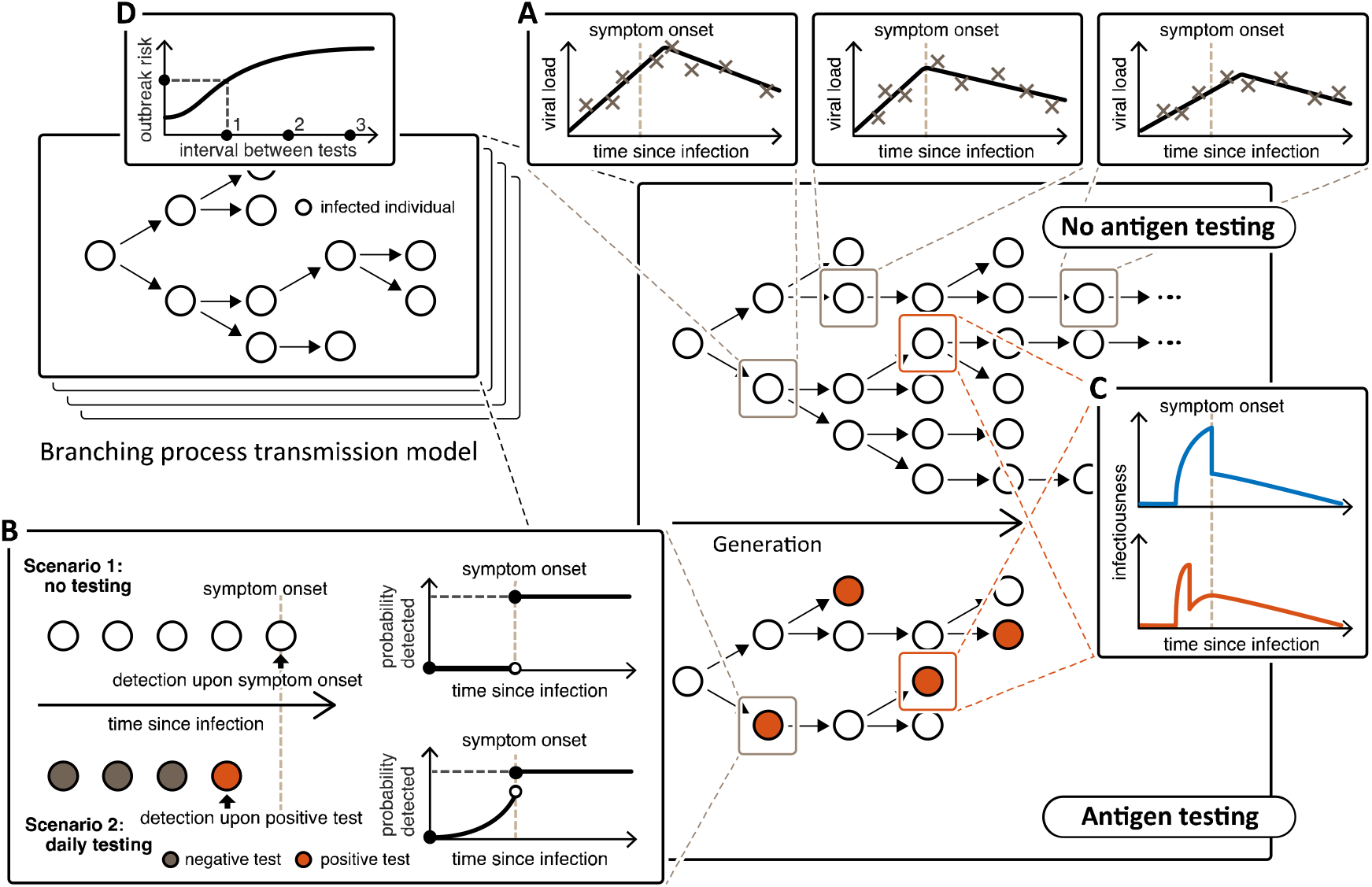
Schematic illustrating our multi-scale approach for calculating the local SARS-CoV-2 outbreak risk, accounting for regular antigen testing. **A** (top right). By fitting a within-host model to individual infection data, the temporal viral load profile(s) of infected individuals can be estimated, potentially accounting for heterogeneity in within-host dynamics between individuals. **B** (bottom left). In the absence of antigen testing, we assumed that infected individuals are detected upon symptom onset (scenario 1). Regular antigen testing of the whole local population may facilitate detection before symptoms (scenario 2), where the viral load profile can be used to estimate the probability of a positive test result. Accounting for the frequency of testing, the probability of detection by each time since infection can be calculated. **C** (middle right). The viral load profile(s) can then be used to estimate the infectiousness profile(s) of infected individuals, accounting for a lower transmission risk following detection, so earlier detection (e.g., when regular antigen testing takes place) leads to a suppressed infectiousness profile. **D** (top left). Assuming a branching process transmission model incorporating the estimated infectiousness profile(s) (the transmission trees next to **C** represent possible model realisations either without or with regular antigen testing), we have analytically derived an equation satisfied by the outbreak risk. The impact of the frequency of antigen testing on the outbreak risk can then be assessed.

### The SARS-CoV-2 local outbreak risk and the impact of regular antigen testing

Using nonlinear mixed effects modelling, we fitted a within-host viral dynamics model (35–42) to data from 521 individuals with infections due to the omicron SARS-CoV-2 variant (43). The temporal viral load profile of infected individuals, using population estimates of within-host model parameters (**Supplementary Table 1**), is shown in **Figure 2A**. Model fits to data from individual hosts are shown in **Supplementary Figure 1**.

**Figure 2.**
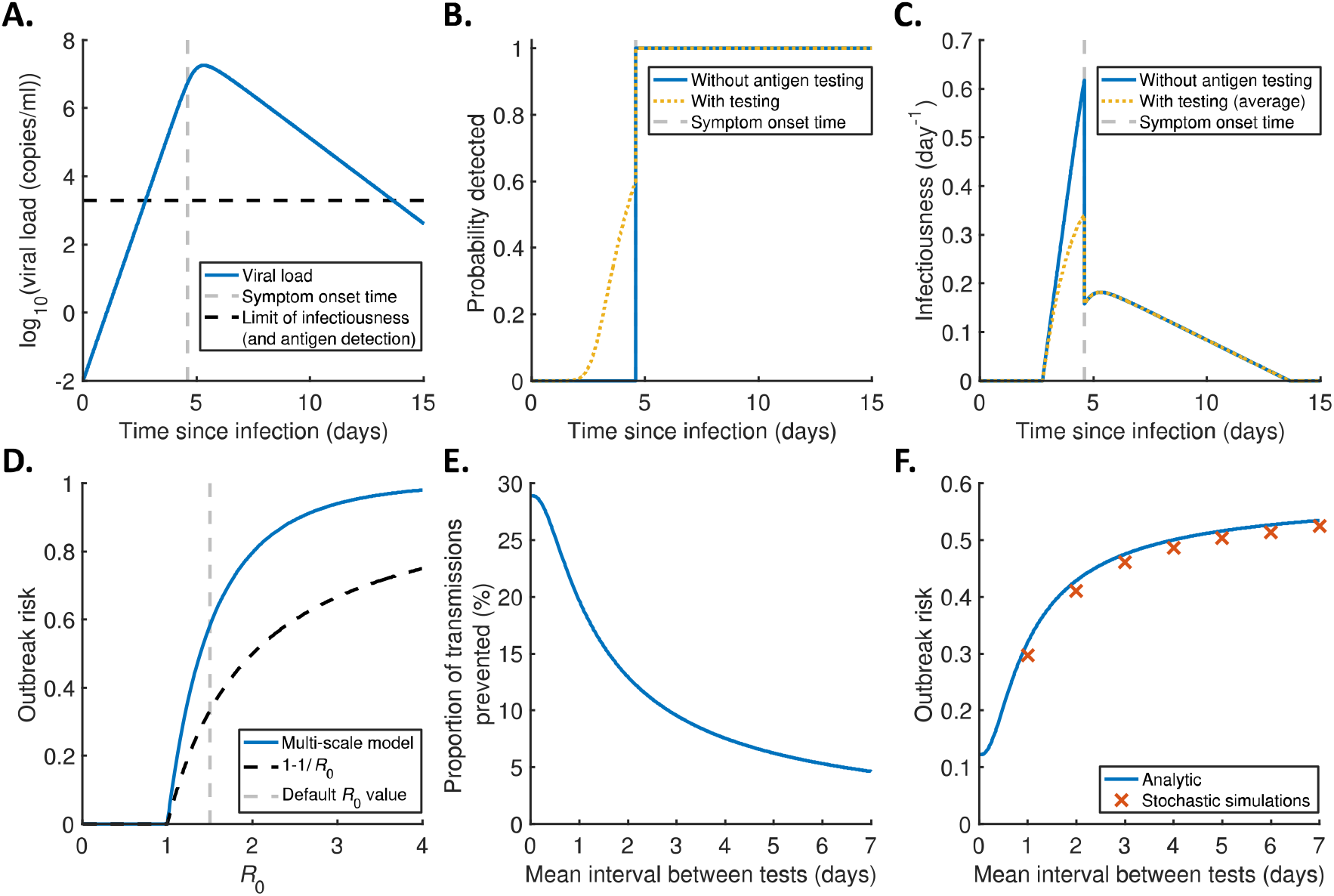
Multi-scale estimation of the SARS-CoV-2 local outbreak risk and analysis of the impact of regular antigen testing. **A**. Viral load profile using population estimates of within-host model parameters (**Supplementary Table 1**). The symptom onset time is shown as a vertical grey dashed line (note that the incubation period was estimated as part of the model fitting procedure), and the assumed viral load threshold for infectiousness and antigen detection is shown as a horizontal black dashed line. Note that we assumed a measurement error affecting antigen test outcomes, leading to the possibility of a positive antigen test with true viral load below this threshold, and vice versa. **B**. Probability of detection by each time since infection, both without regular antigen testing (blue) and with testing every two days (orange dotted). **C**. Infectiousness profiles in the two scenarios, averaging over exact detection times of different individuals in the scenario with antigen testing. **D**. The probability of a major outbreak without antigen testing for different values of the basic reproduction number for local transmissions, *R*_0_, comparing our multi-scale approach (blue) with the commonly used formula, 1− 1/ *R*_0_ (whenever *R*_0_ > 1; black dashed). **E**. The proportion of transmissions prevented from each infected individual by regular antigen testing (compared to a scenario where infected individuals are only detected upon symptom onset), 1 − *R*_0,eff_/*R*_O_ (where *R*_0,eff_ is the basic reproduction number accounting for testing), for different values of the (mean) interval between tests. **F**. The outbreak risk for different values of the (mean) interval between tests when *R*_0_ = 1.5 in the absence of testing (results for other values of *R*_0_ are shown in **Figure 3C**), comparing our analytic multi-scale approach (blue) with estimates obtained using a discrete-time stochastic outbreak simulation model (see **Supplementary Figure 2**; red crosses).

For simplicity, we initially demonstrated our multi-scale approach for estimating the outbreak risk under the assumption of homogeneous within-host dynamics (heterogeneous within-host dynamics are considered in **Figure 4**). First, we used the viral load profile in **Figure 2A** to estimate the probability of detection by each time since infection (**Figure 2B**). We considered scenarios both without regular population antigen testing (so that infected individuals are only detected upon symptom onset), and with regular antigen testing (with the detection probability accounting for randomness in test timing and outcome). We then estimated the (expected) infectiousness profile in each scenario, accounting for a reduction in the transmission risk following detection, and averaging over different possible detection times in the scenario with regular antigen testing (**Figure 2C**).

Outbreak risk estimates in the absence of regular antigen testing, obtained using either our multi-scale approach (using Eq. (1)) or the commonly used population-level estimate (which neglects within-host dynamics), 1 − 1/*R*_0_, are shown for a range of values of *R*_0_ in **Figure 2D** (throughout this article, we always use *R*_0_ to denote the basic reproduction number without regular antigen testing; we denote the reproduction number at the start of the outbreak but accounting for testing (if carried out) by *R*_0,eff_). As would be expected, the outbreak risk increases with *R*_0_, while our multi-scale method generally gives an outbreak risk higher than the standard population-level estimate (when *R*_0_ ≥1).

We then used our multi-scale approach to explore the impact of regular antigen testing on the outbreak risk. First, we estimated the proportion of transmissions prevented from each infected individual, compared to a scenario in which infected individuals are only detected upon symptom onset, under different frequencies of testing (**Figure 2E**). We then calculated the outbreak risk in each case (**Figure 2F**), assuming *R*_0_ = 1.5 in the absence of testing (different values are considered in **Figure 3**). Daily testing was here found to prevent 20% of transmissions (**Figure 2E**), leading to an outbreak risk of 0.32 (**Figure 2F**), which is 45% lower than the corresponding outbreak risk without testing (0.58). In comparison, testing every two days was found to prevent only 13% of transmissions, giving an outbreak risk of 0.43.

**Figure 3.**
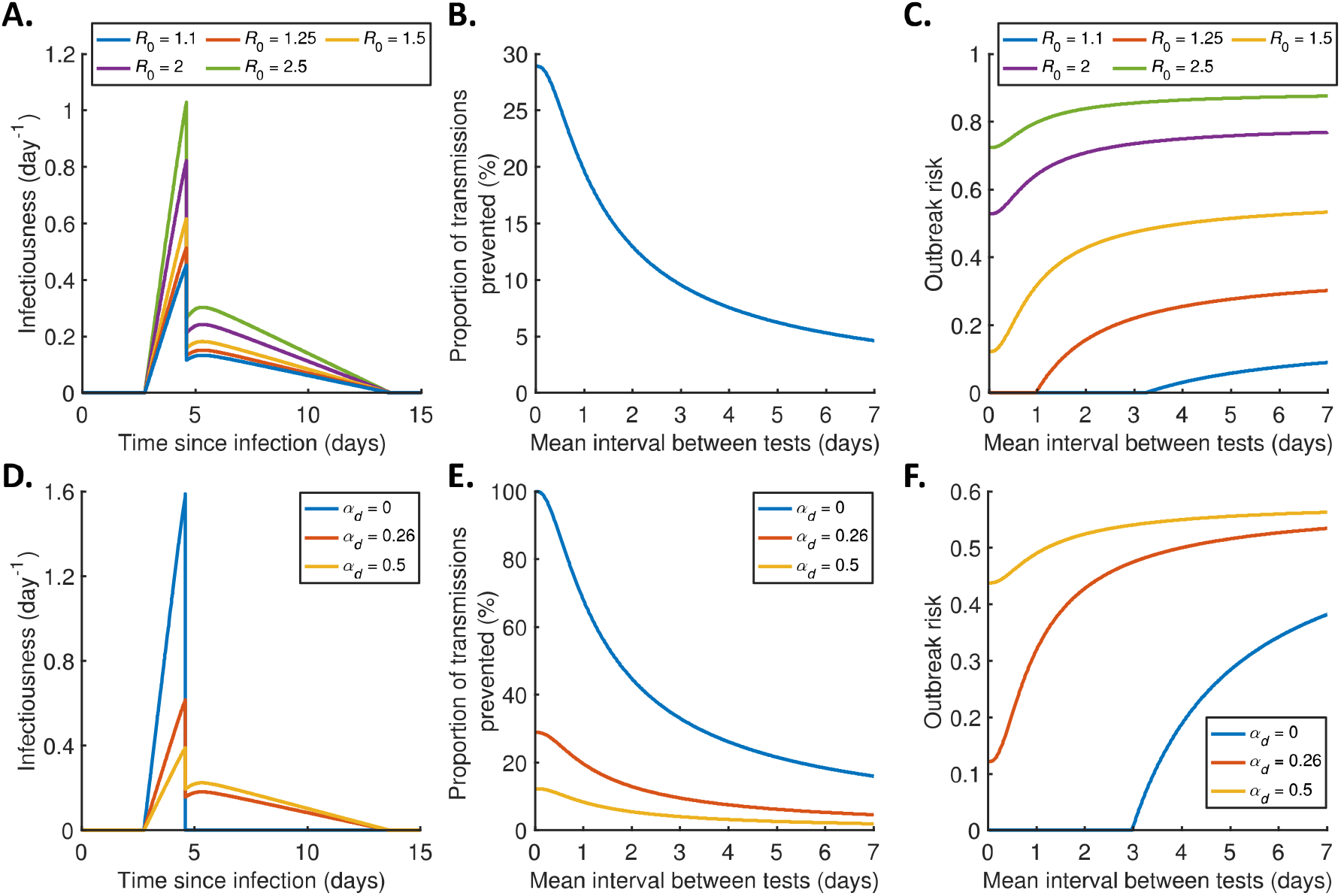
Effect of the local reproduction number and the extent of transmission following detection on the outbreak risk under regular antigen testing. **A**. Infectiousness profiles without regular antigen testing, assuming a basic reproduction number for local transmissions (in the absence of testing) of *R*_0_ = 1.1 (blue), 1.25 (red), 1.5 (orange), 2 (purple), or 2.5 (green). **B**. The proportion of transmissions prevented from each infected individual by regular antigen testing (which is independent of *R*_0_). **C**. The outbreak risk for different values of the (mean) interval between antigen tests, plotted for each *R*_0_ value. **D**. Infectiousness profiles without regular antigen testing, assuming the relative infectiousness of a detected host (compared to an undetected individual with the same viral load) is *α*_*d*_ =0 (blue), 0.26 (red; the value used elsewhere in our analyses (44)), or 0.5 (yellow), with *R*_0_ = 1.5 in all cases. Under these *α*_*d*_ values, the proportions of presymptomatic transmissions (without regular antigen testing) are 100%, 39% and 25%, respectively. **E**. The proportion of transmissions prevented from each infected individual by regular antigen testing, for different values of the (mean) interval between tests, plotted for each *α*_*d*_ value. **F**. The outbreak risk for different values of the (mean) interval between tests, plotted for each *α*_*d*_ value.

To verify our results, we also used a discrete-time, individual-based, stochastic outbreak simulation model to estimate the outbreak risk (**Supplementary Figure 2**). There was relatively close agreement between estimates of the outbreak risk between our analytic approach (blue line in **Figure 2F**) and the stochastic simulations (red crosses). In **Supplementary Figure 3**, we considered the sensitivity of our results to details of how we implemented antigen testing in our multi-scale framework. We found a lower outbreak risk under frequent antigen testing (i.e., antigen testing had a greater impact on the outbreak risk) when we assumed a constant interval between antigen tests, compared to our baseline assumption of a constant rate of testing (i.e., an exponentially distributed interval between tests, which was more straightforward to implement in our analytic approach; **Supplementary Figure 3B**).

### Effect of the local reproduction number and the extent of transmission following detection

In **Figure 2F**, we considered the outbreak risk under antigen testing for a single value of the basic reproduction number for local transmissions (in the absence of testing), *R*_0_ = 1.5. However, even for SARS-CoV-2, the *R*_0_ value for local outbreaks is likely to vary between time periods and local populations because of factors including contact rates, viral evolution and existing immunity levels. Equivalent results to those in **Figure 2F** for different *R*_0_ values are therefore shown in **Figure 3C**. At *R*_0_ values of 1.25 or below, we found daily antigen testing to be sufficient to reduce the outbreak risk to zero (by reducing the reproduction number accounting for testing, *R*_0,eff_, to below one), whereas the estimated outbreak risk remains high even with frequent antigen testing for large *R*_0_ values.

We also explored the effect on our results of the relative transmission risk of detected individuals, *α*_*d*_ (**Figure 3DEF**), with a lower *α*_*d*_ value corresponding to a higher proportion of presymptomatic transmissions. Whereas in most of our analyses we assumed a small, but positive, *α*_*d*_ value (reflecting that, for example, some household transmission may occur following detection) (44), the blue curves in **Figure 3DEF** represent a scenario in which *α*_*d*_ = 0. This may be relevant to specific populations, such as workplaces, in which it may be possible to completely isolate detected cases from the remainder of the population. In the scenario of no transmission from detected individuals, antigen testing at arbitrarily high frequency can theoretically prevent all transmissions that would otherwise occur (whereas in the remainder of our analyses, only some proportion of presymptomatic transmissions can be prevented), with testing every three days here being sufficient to reduce the outbreak risk to zero (when *R*_0_ = 1.5).

### Effect of heterogeneous within-host dynamics and asymptomatic infections

In order to present our multi-scale approach for calculating the outbreak risk in a straightforward setting we have, up to this point, considered a scenario of identical within-host viral dynamics for all infected individuals. However, in reality, within-host dynamics differ between individuals. Our mixed effects within-host model fitting approach has the advantage of facilitating estimation of the extent of heterogeneity in within-host model parameters. We therefore conducted an analysis in which we accounted for such heterogeneity when calculating the localised SARS-CoV-2 outbreak risk (**Figure 4**), using the generalised outbreak risk formulation in Eq. (2).

**Figure 4.**
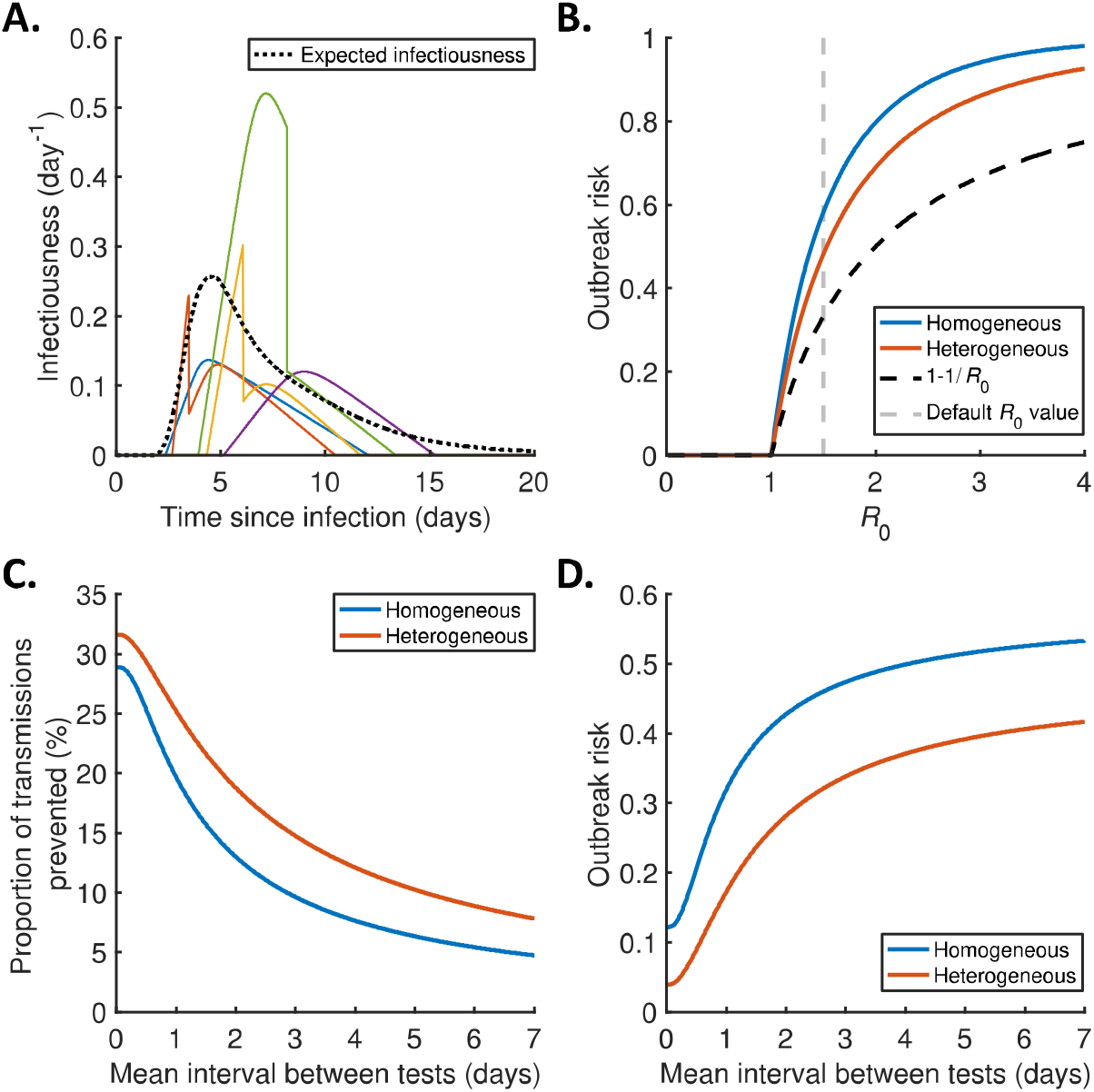
Effect of heterogeneity in within-host dynamics on the outbreak risk under regular antigen testing. **A**. Example simulated infectiousness profiles for five infected individuals in the absence of regular antigen testing, obtained by sampling within-host model parameters using the estimates of fixed and random effects in **Supplementary Table 1** and **Supplementary Table 2**, respectively. The expected infectiousness profile (obtained by averaging the individual infectiousness profiles of a large number of infected individuals) is shown as a black dotted curve. **B**. The probability of a major outbreak without antigen testing for different values of the basic reproduction number, *R*_0_, comparing our multi-scale approach, either assuming homogeneous within-host dynamics (blue) or accounting for heterogeneity (red), and the commonly used formula, 1 − 1/*R*_0_ (black dashed). **C**. The proportion of transmissions per infected individual prevented by regular antigen testing, for different values of the (mean) interval between tests, plotted for the models with homogeneous (blue) and heterogeneous (red) within-host dynamics. **D**. The outbreak risk for different values of the (mean) interval between tests, for the same scenarios as in panel **C**.

We found that accounting for heterogeneity in within-host dynamics leads to a slightly smaller outbreak risk in the absence of regular antigen testing than in **Figure 2** (**Figure 4B**). The model with heterogeneous within-host dynamics also gives a higher proportion of transmissions prevented by regular antigen testing compared to the homogeneous model (for each testing frequency considered; **Figure 4C**), contributing to a greater difference in outbreak risk between the two models with regular antigen testing than without (**Figure 4D**). For example, the outbreak risk when *R*_0_ = 1.5 is 0.48 for the heterogeneous model without testing (0.58 for the homogeneous model), and 0.17 with daily testing (0.32).

We also used the same outbreak risk formulation (Eq. (2)) to account for the possibility of entirely asymptomatic infections (i.e., some individuals remaining without symptoms throughout infection; **Figure 5**). In scenarios with a higher proportion of total transmissions generated by entirely asymptomatic infected hosts, the proportion of transmissions prevented by regular antigen testing was found to be higher (**Figure 5B**). This is because we assumed that asymptomatic hosts remain undetected throughout infection when antigen testing is not carried out, leading to a greater impact of antigen testing on asymptomatic transmissions than on those from individuals who develop symptoms. This effect is likely responsible for a lower outbreak risk at higher proportions of asymptomatic transmissions when antigen testing takes place frequently (**Figure 5C**). We note that an assumption that 0% of transmissions are generated by the asymptomatic infected individuals in the population (blue curve in **Figure 5C**) is different to assuming that there are no asymptomatic infected individuals at all (black dashed curve). For example, in the former case, the outbreak risk will be zero whenever the primary infected individual is asymptomatic, whereas in the latter case the primary infected individual will not remain asymptomatic throughout infection.

**Figure 5.**
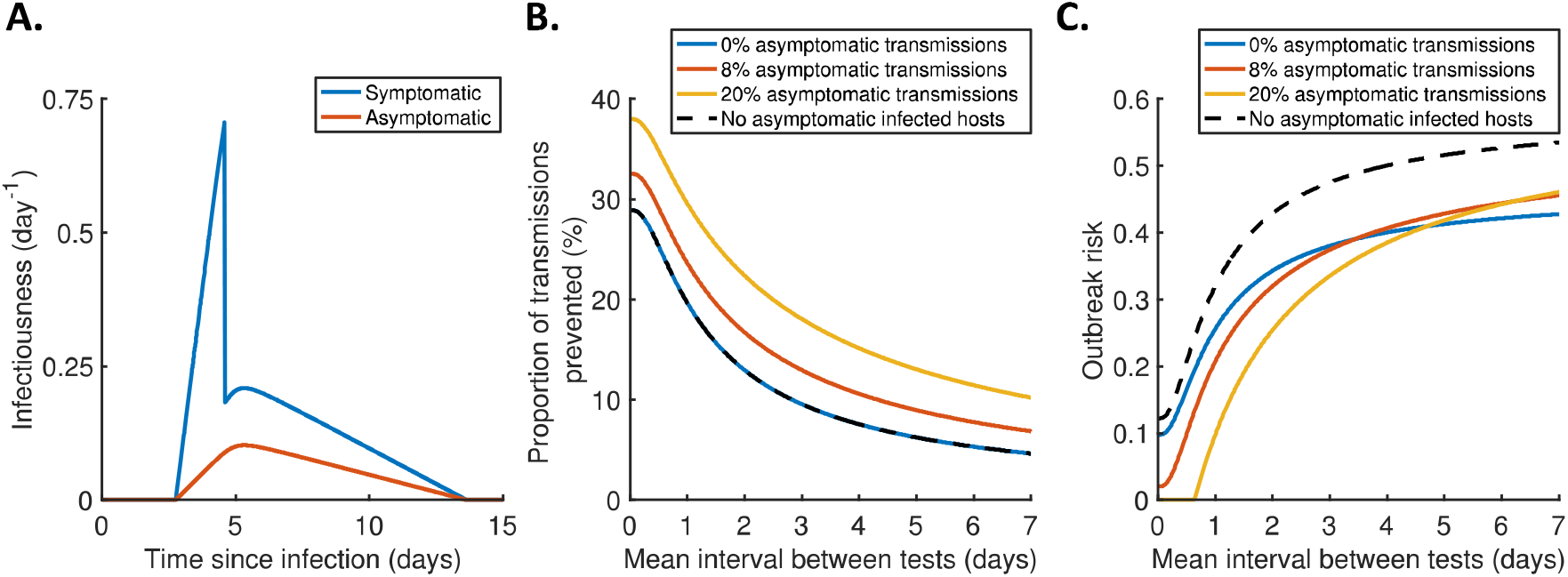
Effect of asymptomatic infections on the outbreak risk under regular antigen testing. **A**. Infectiousness profiles for infected individuals who develop symptoms (blue) and those who remain asymptomatic throughout infection (red), in the absence of regular antigen testing, assuming an overall basic reproduction number, *R*_0_ = 1.5, and that entirely asymptomatic hosts represent 20% of infected hosts and generate 8% of transmissions (45). **B**. The proportion of transmissions per infected individual prevented by regular antigen testing, for different values of the (mean) interval between tests, assuming entirely asymptomatic infected hosts represent 20% of all infected hosts and generate 0% (blue), 8% (red) or 20% (orange) of transmissions in the absence of antigen testing, and assuming no asymptomatic infected hosts (black dotted). **C**. The outbreak risk for different values of the (mean) interval between tests, for the same scenarios as in panel **B**.

### Delayed and/or time-limited antigen testing

In most of our analyses of the effect of regular antigen testing on the local SARS-CoV-2 outbreak risk, we focussed on a scenario in which testing is in place at the time of virus introduction and continues indefinitely. However, we also generalised our analytic outbreak risk derivation to scenarios where the infectiousness profile is calendar time-dependent (**Supplementary Note 4**). This enabled us to explore how the effectiveness of antigen testing is reduced if testing is introduced reactively following the first infection occurring (**Supplementary Figure 4A**), and/or continues for only a limited time (**Supplementary Figure 4B**). We also conducted an analysis in which we assumed a specified total number of tests to be available to each individual (on average), and explored the optimal spacing of these tests to minimise the outbreak risk (for example, 10 tests could be taken daily over 10 days, or once every two days over 20 days; **Supplementary Figure 4C**), assuming reactive testing is introduced when the first infected individual develops symptoms. These analyses therefore demonstrate how antigen testing strategies can be optimised in settings with limited testing resources.

## Discussion

A key challenge for public health policy advisors is estimating the risk that infectious disease cases introduced into a population will lead to a major local outbreak. If the local outbreak risk can be calculated in populations with different characteristics, this will enable limited surveillance and control resources to be targeted effectively. In this article, we have presented a novel modelling framework for estimating the local outbreak risk accounting for within-host viral dynamics.

To demonstrate our multi-scale approach in a concrete setting, we focussed on the risk of local SARS-CoV-2 outbreaks. We used nonlinear mixed effects modelling to fit a within-host model that has been used extensively to model SARS-CoV-2 viral dynamics (35–42) to data from 521 individuals with omicron variant infections (43). The nonlinear mixed effects approach enabled us to quantify the variability in within-host dynamics between individuals which, in turn, could be used to characterise heterogeneity in individual infectiousness profiles (describing how the transmission risk varies during each infection). We then calculated the local outbreak risk based on these data, assuming either homogeneous or heterogeneous within-host dynamics, before testing the effect of regular antigen testing. We found that regular antigen testing can mitigate, but not necessarily eliminate, the outbreak risk, depending on the frequency of testing (for example, in **Figure 2** we estimated an outbreak risk of 0.58 without testing, 0.43 with testing every two days, and 0.32 with daily testing) and local transmission characteristics.

Regular antigen testing is an example of an intervention that can be modelled in greater detail using a multi-scale approach than is possible using a simpler population-level model. This is because both the probability of a positive test result, and the impact of detection on transmission, are likely to depend on the viral load profile, with the timing of testing important for determining the exact outcome. Previous studies have used multi-scale models to analyse the effectiveness of antigen testing for controlling an ongoing outbreak (26–28), but none of those studies considered the impact of antigen testing on the outbreak risk.

Antigen testing was carried out at large scale in countries including the UK (46) earlier in the COVID-19 pandemic, but (similarly to other non-pharmaceutical interventions) has become less commonplace following the roll-out of vaccinations. However, local outbreak prevention remains important in some specific populations in the era of living with COVID-19, for example in care homes due to a high proportion of vulnerable individuals, and our analyses of antigen testing have ongoing relevance to such populations (the UK government continues to provide free tests to care homes (47)). Furthermore, while we used within-host data for the omicron SARS-CoV-2 variant, our methodology and qualitative findings will be applicable if a future SARS-CoV-2 variant, or other viral pathogen, necessitates wider use of non-pharmaceutical interventions.

In the absence of antigen testing, accounting for heterogeneity in within-host dynamics between different hosts generally gave rise to a lower outbreak risk estimate compared to that obtained under the assumption of homogeneous within-host dynamics, while the estimates using both versions of our multi-scale approach were higher than a commonly used outbreak risk estimate that does not account for within-host dynamics (13–19) (**Figure 4B**). These results are consistent with previous comparisons of the outbreak risk between models with different infectious period distributions (20) or offspring distributions (48), although previous studies did not consider variations in infectiousness during infection. More variability in the total number of transmissions generated by different individuals typically leads to a lower outbreak risk since, for example, the probability of the primary infected individual generating no transmissions will then be higher. We also found a greater impact of antigen testing on transmission with heterogeneous than homogeneous within-host dynamics (**Figure 4C**), which contributed to a bigger difference in outbreak risk estimates between the heterogeneous and homogeneous models when antigen testing is carried out (**Figure 4D**) than without testing.

Our results highlight that transmission characteristics depending on both the virus and local population under consideration are important in determining the outbreak risk and impact of antigen testing. In settings where the reproduction number for local transmissions is high (e.g., in high-contact environments, or due to a new viral variant or waning immunity), the outbreak risk may remain high even with a high testing frequency, so that mitigations in addition to antigen testing would be required to substantially reduce the risk. Conversely, we found antigen testing to be more effective when a high proportion of transmissions are presymptomatic, such as in schools and workplaces (provided symptomatic individuals are instructed to stay at home). This is because population-wide testing enables infected individuals to be detected before symptoms, thus preventing presymptomatic transmissions that would otherwise have occurred. Similarly, when we accounted for entirely asymptomatic infections (**Figure 5**), we found a lower outbreak risk under daily testing when a higher proportion of transmissions are generated by asymptomatic infectors.

Like any modelling study, our analyses involved assumptions and simplifications. We assumed that infectiousness scales with the logarithm of the viral load (26,30), with a reduction in transmission risk upon detection (due to detected individuals staying at home and/or isolating) (44,49,50). However, more complex within-host models or relationships between viral load and infectiousness, or a delay between detection and isolation, would be straightforward to implement in our multi-scale modelling framework. We also assumed equal viral load thresholds for infectiousness and for antigen test positivity, but this assumption could be relaxed to explore how the outbreak risk under regular antigen testing depends on test sensitivity, which may vary between tests developed by different manufacturers (51). While our focus here was rapid antigen testing, future work may compare the effectiveness of antigen and PCR testing for reducing the outbreak risk, particularly considering a trade-off between test sensitivity and turn-around time that has previously been explored in the context of controlling an ongoing outbreak (28,30).

Our multi-scale approach for estimating the outbreak risk, accounting for heterogeneous within-host viral dynamics, could be extended in numerous directions. We considered a scenario involving a single infected individual arriving in a host population early in their course of infection. However, it would be straightforward to consider possibilities such as the primary infected individual entering the population later in infection, and/or multiple infectious importations occurring. A future study may also relate heterogeneity in within-host dynamics to specific characteristics such as age, enabling the outbreak risk to be compared between local populations with different structures. Other forms of heterogeneity, such as in susceptibility and/or contact rates, could also be considered. Finally, going forwards, we plan to use the mathematical results (**Supplementary Note 4**) underlying our analysis of reactively introduced antigen testing (**Supplementary Figure 4)** to explore temporal changes in the SARS-CoV-2 local outbreak risk, combining our multi-scale approach here with previous work incorporating time-dependent susceptibility into outbreak risk estimates (11) (for example, a booster vaccination campaign followed by waning immunity could be considered).

In summary, we have developed a multi-scale modelling framework in which within-host viral dynamics models can be used to inform estimates of the risk of infectious disease outbreaks and to analyse the impact of pre-emptive control. Applying our approach to estimate the risk of local SARS-CoV-2 outbreaks, we found that regular antigen testing of the local population can reduce, but not eliminate, the outbreak risk, depending on the frequency of testing as well as transmission characteristics that are likely to vary temporally and between different populations. Additionally, we found that it may be important to account for details such as asymptomatic infection and heterogeneity in within-host dynamics to assess the effectiveness of antigen testing accurately. We hope that this research will help to guide pre-emptive control and mitigate the risk of outbreaks due to a range of viruses.

## Methods

### Study data

We analysed published viral load data from 521 individuals with symptomatic infections due to the omicron SARS-CoV-2 variant (43). For each individual in the dataset, the results and timing (relative to a recorded symptom onset date, including some tests carried out prior to symptom onset) of at least three RT-qPCR tests were available. The median number of tests per individual was 15. Viral load values (converted from Ct values) were recorded for positive tests.

### Within-host model and parameter estimation

We used a simple within-host model of SARS-CoV-2 viral dynamics (35–42), given by

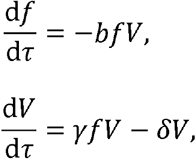

where *f*(*τ*) and *V*(*τ*) denote, respectively, the proportion of uninfected target cells (so that *f*(0) = 1) and viral load at time since infection *τ*. The parameters *b, γ* and *δ* are the rate constant for virus infection, the maximum rate constant for viral replication, and the death rate of infected cells, respectively.

We estimated the parameters *b, γ* and *δ*, in addition to the incubation period, *τ*_inc_, by fitting the model to the viral load data using a nonlinear mixed effects modelling approach (amounting to a partial pooling of the data from each individual). Specifically, the value of the parameter vector, *θ*_*j*_ = (*b*_*j*_, *γ*_*j*_, *δ*_*j*_, *τ*_inc,*j*_), for a given individual, *j*, was assumed to be of the form 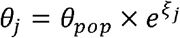 (where the operations are applied element-wise). Here, *θ*_*pop*_ is a fixed effect (referred to as the population parameter value), and *ξ*_*j*_ is a random effect, assumed to be normally distributed with mean zero and covariance matrix *Ω*. For simplicity, we assumed the random effects for different parameters to be independent, with standard deviations *ω*_*b*_, *ω*_*γ*_, *ω*_*δ*_ and 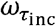 (i.e., 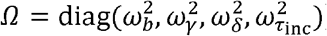).

In the parameter fitting procedure, we estimated both the fixed effects (**Supplementary Table 1**; note that the subscript *pop* is suppressed) and the standard deviations of the random effects (**Supplementary Table 2**). In other words, we characterised both the population (median) values of model parameters, as well as variability in those parameters between individuals. The measurement error, *α*, in recorded values of the log viral load, was also estimated. These parameters were estimated by using the Stochastic Approximation of the Expectation-Maximization (SAEM) algorithm (52,53) to obtain the parameter values that maximise the likelihood of the recorded viral load data. We accounted for left censoring of viral load data (i.e., a negative test result occurring at viral loads below the detection limit of 10^2.66^ copies/ml) in the likelihood. Initial values of estimated parameters were changed multiple times to confirm the robustness of parameter estimation and ensure a global maximum of the likelihood was obtained. Additionally, we calculated best-fit estimates (Empirical Bayes Estimates (53)) of within-host model parameters for each individual host (**Supplementary Figure 1**). Fitting was implemented in MONOLIX version 2019R2 (53).

### Detection model

We assumed that infected individuals could be detected in two possible ways:

1. By returning a positive antigen test.
2. By developing symptoms (we assumed previously undetected hosts to be detected immediately upon symptom onset).

Supposing that an infected individual conducts an antigen test when their instantaneous viral load is *V*, we assumed (similarly to previous work (40,42)) that a positive test result occurs with probability 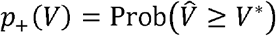. Here, 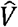 represents a measured viral load, assumed to be normally distributed on the log scale (independently of previous viral load measurements) such that 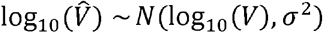; *V*\* is the detection limit (the choice of *V*\* is described in **Supplementary Table 1**); and the value of the measurement error level, *σ*, was assumed to be equal to the corresponding quantity that we estimated for PCR testing (**Supplementary Table 1**). In other words, a positive test result was assumed to occur whenever the measured viral load exceeds the detection limit.

In most of our analyses of regular antigen testing, we assumed an exponentially distributed interval between successive tests, with mean *T* (i.e.,a constant rate of testing; the alternative scenario of a constant interval between tests is considered in **Supplementary Figure 3B**). Under this assumption, the probability of an infected individual being detected by time since infection *τ* is given by

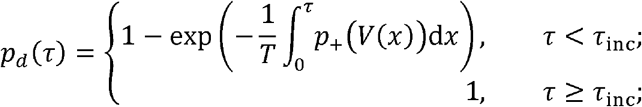

where *V*(*x*) represents the viral load of the specified individual at time since infection *x*, and *τ*_inc_ their incubation period (*τ*_inc_ can be taken to be infinite to represent an entirely asymptomatic infection). This expression is derived in **Supplementary Note 1**.

### Infectiousness model

The infectiousness profile of an undetected host, *β*_*u*_(*τ*), at each time since infection, *τ*, was assumed to depend on their viral load, *V*(*τ*), according to a prescribed functional relationship. Specifically, we assumed (26,30)

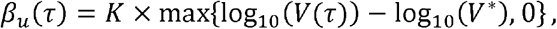

so that only individuals with a viral load exceeding *V*\* are infectious. We assumed this infectiousness limit to be equal to the detection limit for antigen testing (i.e., in the absence of measurement errors, infected individuals will return a positive antigen test if and only if they are infectious at the time of testing). The choice of the scaling factor, *K*, is described below.

We assumed the effective infectiousness (accounting for behavioural factors) of a detected individual at time since infection *τ* (where *τ* exceeds the time of detection) to be a factor *α*_*d*_ times *β*_*u*_(*τ*) (the choice of *α*_*d*_, which lies between zero and one, is described in **Supplementary Table 1**). In the absence of regular antigen testing, the overall individual infectiousness profile is then

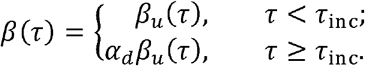

When regular antigen testing takes place, supposing the individual under consideration has been detected by time since infection *τ* with probability *p*_*d*_(*τ*), then their expected infectiousness at time since infection *τ* (accounting for different possible detection times) is

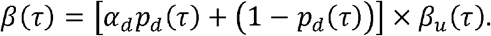

The scaling factor, *K*, in the expression for *β*_*u*_(*τ*), was chosen to obtain a specified value of the basic reproduction number, *R*_0_, in the absence of regular antigen testing (except where otherwise specified, we took the default value *R*_0_ = 1.5). Specifically, if the expected infectiousness profile (averaging over individual infectiousness profiles if the population is heterogeneous) is 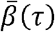, then we have (see **Supplementary Note 2** in detail)

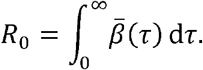

### Outbreak risk

Here, we describe our approach for calculating the (local) outbreak risk (the probability that a major outbreak results from a single newly infected individual being introduced into an otherwise uninfected population) under the within-host, detection and infectiousness models described above. A benefit of our approach is that we have derived equations satisfied by the (local) outbreak risk analytically under a branching process transmission model, assuming either homogeneous or heterogeneous within-host dynamics between different infected individuals. These equations are described below; derivations are given in **Supplementary Note 2**. The equations were solved numerically, avoiding the need to run large numbers of stochastic model simulations to estimate the local outbreak risk. However, we also verified our analytically derived outbreak risk estimates against simulations of a discrete-time, individual-based, stochastic epidemic model in **Figure 2F** (see **Supplementary Figure 2** and **Supplementary Note 5** for details).

#### Homogeneous population model

First, we considered a simplified scenario in which each member of the population is assumed to follow the same infectiousness profile, *β*(*τ*) In this case, an analytic argument gives the following implicit equation for the outbreak risk:

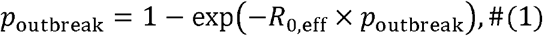

where the largest solution between 0 and 1 should be taken. Here, *R*_0,eff_ is the reproduction number at the start of the outbreak, accounting for regular antigen testing (assumed to be in place at the time of pathogen introduction) if carried out, while we use *R*_0_ to refer specifically to the basic reproduction number in the absence of regular antigen testing. This equation has previously been derived in the special case of constant infectiousness during a fixed infectious period (18), but we show in **Supplementary Note 2** that this equation is valid for any *β*(*τ*) (provided there is no heterogeneity in infectiousness). We also emphasise that while this equation only depends on *R*_0,eff_, even in this simplified scenario our multi-scale approach enables detailed analysis of how interventions such as antigen testing affect *R*_0,eff_ and therefore the outbreak risk, which cannot otherwise be captured easily.

We solved Eq. (1) numerically under the parameter values in **Supplementary Table 1** (in particular, using population estimates of the within-host parameters *b, γ, δ* and *τ*_inc_) in order to estimate *p*_outbreak_ in the absence of regular antigen testing. Then, we explored the effect of antigen testing on *p*_outbreak_, for simplicity averaging over the exact detection times of different individuals in most of our analyses. Of note, in **Supplementary Figure 3A**, we found that explicitly accounting for heterogeneity in detection times had a very small effect on the *p*_outbreak_ estimates.

#### Heterogeneous population model

We also conducted an analysis in which we accounted for heterogeneity in within-host dynamics between different individuals. In **Supplementary Note 2**, we show that if there are *n* population subgroups, with each infected individual in group *j* assumed to follow infectiousness profile *β*_*j*_(*τ*), then the outbreak risk is the largest solution between 0 and 1 of

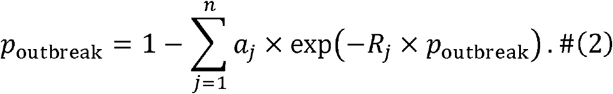

Here, *a*_*j*_ represents the proportion of new infections that are in group *j* (at the start of the epidemic and neglecting heterogeneity in contact rates between groups), which simply corresponds to the proportion of the population in group *j* if there is no difference in susceptibility between groups, while *R*_*j*_ gives the expected number of transmissions generated by an infected host in group *j* over the course of infection (at the start of the outbreak). We note that Eq. (2) includes as special cases most previous outbreak risk estimates based on branching process approximations of compartmental epidemic models (see **Supplementary Note 3**).

To account for heterogeneous within-host dynamics, we used the estimated fixed (**Supplementary Table 1**) and random (**Supplementary Table 2**) effects to generate synthetic viral load profiles and incubation periods for *n* = 10,000 infected individuals. The infectiousness profile of each individual was obtained (averaging over possible detection times when analysing regular antigen testing), and then Eq. (2) was used to estimate the outbreak risk (taking *a*_*j*_ = 1/*n* for each *j*).

We also used Eq. (2) when we accounted for entirely asymptomatic infections (**Figure 5**). In **Figure 5**, we assumed that a proportion, *a*_1_ = 0.8, of infected individuals develop symptoms, with the remaining proportion, *a*_2_ = 0.2, remaining asymptomatic throughout infection (45) (i.e., we took *n* = 2 in Eq. (2)). For simplicity, we assumed no difference in within-host model parameters between entirely asymptomatic hosts and those who develop symptoms, but instead considered different possible values of the proportion of all transmissions arising from entirely asymptomatic infectors (in the absence of regular antigen testing), given by

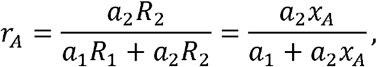

where *x*_*A*_ = *R*_2_/*R*_1_ represents the relative overall transmissibility of asymptomatic infected hosts (where *R*_1_ and *R*_2_ represent the expected total number of transmissions generated by each infected host who develops symptoms and by each entirely asymptomatic infected host, respectively). Specifically, we considered *r*_*A*_ values of 0, 0.08 and 0.2, which were obtained by scaling the infectiousness profiles of entirely asymptomatic hosts to obtain corresponding *x*_*A*_ values of 0, 0.35 (the value obtained by (45)) and 1, respectively.

Finally, we also used Eq. (2) when we accounted for variability in the exact detection times of different infected individuals under regular antigen testing, assuming either an exponentially distributed interval between tests (**Supplementary Figure 3A**) or a fixed interval (**Supplementary Figure 3B**; in this case assuming each infection time to be uniformly distributed between two testing times). In both cases, we sampled the detection times of *n* = 10,000 hosts.

### Delayed and/or time-limited regular antigen testing

In most of our analyses, we focussed on a scenario in which regular antigen testing is already in place at the time of pathogen introduction and continues indefinitely. However, we also considered scenarios in which testing is introduced reactively after an infection occurs within the local population, and/or testing is only carried out for a limited time period (**Supplementary Figure 4**). An equation for the outbreak risk in this scenario is derived in **Supplementary Note 4**.

## Data Availability

The viral load data used in our analyses are available at https://doi.org/10.7554/eLife.81849 (Hay et al., eLife, 2022). Code to reproduce our results is available at https://github.com/will-s-hart/multiscale-outbreak-risk.

https://github.com/will-s-hart/multiscale-outbreak-risk

## Acknowledgements

We would like to thank Philip Maini for helpful comments on the manuscript. W.S.H. acknowledges funding by the Engineering and Physical Sciences Research Council via a Doctoral Prize (grant number EP/W524311/1) and by the Japan Society for the Promotion of Science via an International Research Fellowship (short-term Predoctoral Fellowship). This study was supported in part by a Grant-in-Aid for Transformative Research Areas A 22H05215 (to S.I.), JSPS Scientific Research (KAKENHI) B 18H01139 (to S.I.), 16H04845 (to S.I.), Scientific Research in Innovative Areas 20H05042 (to S.I.); AMED CREST 19gm1310002 (to S.I.); AMED Development of Vaccines for the Novel Coronavirus Disease, 21nf0101638s0201 (to S.I.); AMED Japan Program for Infectious Diseases Research and Infrastructure, 20wm0325007h0001 (to S.I.), 20wm0325004s0201 (to S.I.), 20wm0325012s0301 (to S.I.), 20wm0325015s0301 (to S.I.); AMED Research Program on HIV/AIDS 22fk0410052s0401 (to S.I.); AMED Research Program on Emerging and Re-emerging Infectious Diseases 20fk0108140s0801 (to S.I.), 21fk0108428s0301 (to S.I.); AMED Program for Basic and Clinical Research on Hepatitis 21fk0210094 (to S.I.); AMED Program on the Innovative Development and the Application of New Drugs for Hepatitis B 22fk0310504h0501 (to S.I.); AMED Strategic Research Program for Brain Sciences 22wm0425011s0302; JST MIRAI JPMJMI22G1 (to S.I.); Moonshot R&D JPMJMS2021 (to S.I.) and JPMJMS2025 (to S.I.); Shin-Nihon of Advanced Medical Research (to S.I.); SECOM Science and Technology Foundation (to S.I.).

## Author contributions

W.S.H. and S.I. conceptualised the study. All authors developed the methodology. W.S.H. and H.P. wrote the model code and conducted the computational analyses. S.I. and R.N.T. supervised the research. W.S.H. wrote the initial manuscript draft. All authors reviewed and edited the manuscript.

