## Supplementary Information for "Analysis of the risk and pre-emptive control of viral outbreaks accounting for within-host dynamics: SARS-CoV-2 antigen testing as a case study"

[1]Mathematical Institute, University of Oxford, Oxford, OX2 6GG, UK. [2]interdisciplinary Biology Laboratory (iBLab), Division of Natural Science, Graduate School of Science, Nagoya University, Japan. [3]Department of Mathematics, Pusan National University, Busan, South Korea. [4]Department of Science System Simulation, Pukyong National University, Busan, South Korea. [5]Mathematics Institute, University of Warwick, Coventry, CV4 7AL, UK. [6]Zeeman Institute for Systems Biology and Infectious Disease Epidemiology Research (SBIDER), University of Warwick, Coventry, CV4 7AL, UK. [7]Institute of Mathematics for Industry, Kyushu University, Fukuoka, Japan. [8]Institute for the Advanced Study of Human Biology (ASHBi), Kyoto University, Kyoto, Japan. [9]Interdisciplinary Theoretical and Mathematical Sciences Program (iTHEMS), RIKEN, Saitama, Japan. [10]NEXT-Ganken Program, Japanese Foundation for Cancer Research (JFCR), Tokyo, Japan. 13Science Groove Inc., Fukuoka, Japan.

### Supplementary Note 1: Probability of detection

Here, we derive the expression given in the main text for the probability, $p_d(\tau)$, of an infected individual, subject to regular antigen testing, having been detected by time since infection $\tau$. We assume (as in most of our analyses) an exponentially distributed interval of mean $T$ between successive tests (i.e., a constant rate of testing).

First, since we assume that symptomatic hosts are always detected, we have $p_d(\tau) = 1$ for $\tau \geq \tau_{\text{inc}}$, where $\tau_{\text{inc}}$ is the individual's incubation period (which can be taken to be infinite to represent an entirely asymptomatic infection). Now, for $\tau < \tau_{\text{inc}}$, we consider a short time interval $[\tau, \tau + \mathrm{d}\tau]$. The probability that the individual returns a positive antigen test in this interval is given (up to terms of order $\mathrm{d}\tau^2$) by $(1/T)\mathrm{d}\tau \times p_+(V(\tau))$, where $(1/T)\mathrm{d}\tau$ represents the probability of taking a test, and $p_+(V(\tau))$ the probability that the result of a test is positive (which depends on the instantaneous viral load, $V(\tau)$). Conditioning on whether or not a positive test is returned in the interval $[\tau, \tau + \mathrm{d}\tau]$ then gives

$$p_d(\tau + \mathrm{d}\tau) = 1 \times \frac{p_+\big(V(\tau)\big)\mathrm{d}\tau}{T} + p_d(\tau) \times \left(1 - \frac{p_+\big(V(\tau)\big)\mathrm{d}\tau}{T}\right),$$

where the probability of detection by time $(\tau + \mathrm{d}\tau)$ conditional on a positive test in $[\tau, \tau + \mathrm{d}\tau]$ is $1$, and the probability conditional on no positive test in that interval is $p_d(\tau)$.

Rearranging the above equation and taking the limit $\mathrm{d}\tau \to 0$ gives the differential equation,

$$\frac{\mathrm{d}p_d}{\mathrm{d}\tau} = \frac{p_+\big(V(\tau)\big)}{T}\big(1 - p_d(\tau)\big),$$

which can be solved alongside the initial condition $p_d(0) = 0$ to obtain

$$p_d(\tau) = 1 - \exp\left(-\frac{1}{T}\int_0^\tau p_+\big(V(x)\big)\mathrm{d}x\right),$$

for $\tau < \tau_{\text{inc}}$.

### Supplementary Note 2: Derivation of outbreak risk

Below, we derive an analytical expression for the outbreak risk in a heterogeneous population divided into $n$ subgroups (the special case of a homogeneous population is obtained when $n = 1$), between which the infectiousness profile of infected hosts (as well as other factors such as susceptibility) may vary. Specifically, we consider a branching process model in which susceptible depletion is neglected and infection lineages are assumed to be independent. The outbreak risk is then taken to be the probability that epidemic extinction does *not* occur within this branching process framework, following the introduction of a single newly infected host into the population, i.e., the probability that the number of currently infected individuals never reaches zero but instead tends to infinity (note that in reality, the assumptions underlying the branching process model will no longer be valid when the number of infected individuals becomes large).

We suppose that each infected host in group $j$ transmits the pathogen to individuals in group $i$ at total rate $\beta_{i,j}(\tau)$ at time since infection $\tau$ (a specific form of $\beta_{i,j}(\tau)$ is considered later). The expected total number of infections generated in group $i$ by each infected host in group $j$ (over the course of infection) is then

$$R_{i,j} = \int_0^\infty \beta_{i,j}(\tau)\mathrm{d}\tau\,,$$

where the basic reproduction number (accounting for regular antigen testing if in place), $R_{0,\mathrm{eff}}$, is the largest eigenvalue of the matrix with entries $R_{i,j}$ (the next-generation matrix) (1).

Now, we suppose that a single infected individual in group $j$ is introduced into the population at time since infection $\tau$, with the remainder of the population assumed to be uninfected at the time of introduction (and assuming no further external pathogen introductions into the population). Then an expression for the probability of extinction (i.e., the probability that a major outbreak does *not* occur), denoted $q_j(\tau)$, can be derived by conditioning on whether or not the initial infected individual transmits the pathogen (to an individual in any population group) between times since infection $\tau$ and $(\tau + \mathrm{d}\tau)$, to obtain (neglecting the possibility that multiple transmissions occur, which has probability of order $\mathrm{d}\tau^2$)

$$q_j(\tau) = \left(\sum_{i=1}^{n} q_j(\tau + \mathrm{d}\tau) q_i(0) \times \beta_{i,j}(\tau)\mathrm{d}\tau\right) + q_j(\tau + \mathrm{d}\tau) \times \left(1 - \sum_{i=1}^{n} \beta_{i,j}(\tau)\mathrm{d}\tau\right).$$

Here, $\beta_{i,j}(\tau)\mathrm{d}\tau$ gives the probability of a transmission to an individual in group $i$ occurring in this time interval, $q_j(\tau+\mathrm{d}\tau)q_i(0)$ the extinction probability conditional on such a transmission occurring (since infection lineages are assumed to be independent), and $q_j(\tau+\mathrm{d}\tau)$ the extinction probability conditional on no transmissions occurring.

Rearranging the above equation and taking the limit $\mathrm{d}\tau \to 0$ gives the differential equation,

$$\frac{\mathrm{d}q_j}{\mathrm{d}\tau} = q_j(\tau)\sum_{i=1}^{n}\big(1-q_i(0)\big)\beta_{i,j}(\tau),$$

which can be solved alongside the boundary condition $q_j(\infty)=1$ to obtain

$$q_j(\tau) = \exp\left(-\sum_{i=1}^{n}\big(1-q_i(0)\big)\int_{\tau}^{\infty}\beta_{i,j}(x)\mathrm{d}x\right).$$

In particular, we have

$$q_j(0) = \exp\left(-\sum_{i=1}^{n}\big(1-q_i(0)\big)R_{i,j}\right).$$

Now, a relatively general parameterisation is to take $\beta_{i,j}(\tau)=\varepsilon_i\eta_i C_{i,j}\beta_j(\tau)$, so that $R_{i,j}=\varepsilon_i\eta_i C_{i,j}B_j$. Here, $\beta_j(\tau)$ is the infectiousness profile of an infected individual in group $j$, $B_j$ is the total integral of $\beta_j(\tau)$ over all times since infection, $\varepsilon_i$ is the proportion of the population who are in group $i$, $\eta_i$ is the relative susceptibility in group $i$, and $C_{i,j}$ represents the rate of contacts between individuals in groups $i$ and $j$. The above equation can be used to calculate the outbreak risk for general $\beta_{i,j}(\tau)$. However, further analytic progress is possible under the assumption of homogeneous mixing (i.e., when $C_{i,j}$ is independent of $i$ and $j$). Absorbing the value of $C_{i,j}$ into $\beta_{i,j}(\tau)$ (i.e., setting $C_{i,j}=1$), we then have

$$q_j(0) = \exp\left(-B_j\sum_{i=1}^{n}\big(1-q_i(0)\big)\varepsilon_i\eta_i\right). \qquad (\mathrm{S1})$$

In this case, the overall extinction probability following the introduction of a single newly infected individual is

$$q(0) = \frac{1}{\bar{\eta}}\sum_{j=1}^{n}\varepsilon_j\eta_j q_j(0) = \sum_{j=1}^{n}a_j q_j(0), \qquad (\mathrm{S2})$$

where $\bar{\eta} = \sum_{j=1}^{n} \varepsilon_j \eta_j$ gives the mean population susceptibility, and $a_j = \varepsilon_j \eta_j / \bar{\eta}$ gives the proportion of new infections that are in group $j$. Now, Eq. (S1) above can be written as

$$q_j(0) = \exp\left(-\left(1 - q(0)\right)\bar{\eta} B_j\right),$$

and substituting this expression into Eq. (S2) then gives

$$q(0) = \frac{1}{\bar{\eta}} \sum_{j=1}^{n} \varepsilon_j \eta_j \exp\left(-\left(1 - q(0)\right)\bar{\eta} B_j\right) = \sum_{j=1}^{n} a_j \exp\left(-\left(1 - q(0)\right) R_j\right), \qquad (S3)$$

where $R_j = \bar{\eta} B_j$ gives the expected number of transmissions generated by an infected host in group $j$ (over the course of infection).

Finally, the outbreak risk (following the introduction of a single newly infected individual into an otherwise susceptible population), $p_{\mathrm{outbreak}} = 1 - q(0)$, then satisfies

$$p_{\mathrm{outbreak}} = 1 - \sum_{j=1}^{n} a_j \times \exp\left(-R_j \times p_{\mathrm{outbreak}}\right),$$

which is Eq. (2) in the main text. While this equation may have multiple solutions (in particular, $p_{\mathrm{outbreak}} = 0$ is always a solution), by standard theory of hitting probabilities on Markov chains (2), the relevant solution is the largest solution between 0 and 1 (since the relevant solution to Eq. (S3) is the minimal non-negative one). While we focussed on the outbreak risk starting with a single, newly infected, primary case, it would be straightforward to consider an infected individual introduced into the population later in infection, and/or multiple pathogen introductions, in our approach.

We note that the basic reproduction number in this scenario (accounting for regular antigen testing, if carried out) is

$$R_{0,\mathrm{eff}} = \sum_{i=1}^{n} \varepsilon_i \eta_i B_i = \sum_{i=1}^{n} a_i R_i,$$

where the first equality follows since the next-generation matrix is of separable form $R_{i,j} = c_i d_j$ (where here $c_i = \varepsilon_i \eta_i$ and $d_j = B_j$) and therefore has largest eigenvalue $R_{0,\mathrm{eff}} = \sum_{i=1}^{n} c_i d_i$ (1). This can also be written as

$$R_{0,\mathrm{eff}} = \int_0^\infty \bar{\beta}(\tau)\,\mathrm{d}\tau,$$

where $\bar{\beta}(\tau) = \bar{\eta} \sum_{i=1}^{n} a_i \beta_i(\tau)$ gives the expected infectiousness profile, accounting for the relative susceptibility of the population. We also note that while the derivation here includes the possibility of heterogeneous susceptibility between different population sub-groups, we did not consider heterogeneous susceptibility in our numerical analyses.

In the special case of a homogeneous population ($n = 1$), we have

$$p_{\text{outbreak}} = 1 - \exp\left(-R_{0,\text{eff}} \times p_{\text{outbreak}}\right),$$

i.e. we recover Eq. (1) in the main text.

### Supplementary Note 3: Wider applicability of outbreak risk equation accounting for heterogeneity

The result in Eq. (2), while derived in the context of a time-since-infection model in a heterogeneous population, is in fact widely applicable to a range of (branching process) models. Specifically, taking the limit of a continuous distribution of population subgroups in Eq. (2) gives the equation

$$p_{\text{outbreak}} = 1 - \int_{\Theta} a(\theta) \times \exp(-R(\theta) \times p_{\text{outbreak}}) \mathrm{d}\theta. \qquad (S4)$$

Here $\theta \in \Theta$ is a continuous variable (which may be either real-valued or higher-dimensional) indexing population sub-groups and/or possible "types" of infection, $a(\theta)$ is the probability density that a new infection is of type $\theta$, and $R(\theta)$ gives the expected total number of transmissions generated by an infected host with infection type $\theta$. We briefly note that the continuous formulation in Eq. (S4) is applicable to the scenario of heterogeneous within-host dynamics that we considered, but in practice it was easier to calculate the outbreak risk by sampling the within-host dynamics of a large number of hosts and using Eq. (2).

As an example to demonstrate the applicability of Eq. (S4), we consider a branching process approximation of the stochastic SIR compartmental epidemic model. In this case, the possible "types" of infection are indexed by the infectious period, $\theta = t_I \in [0, \infty)$, with $a(\theta) = \mu \exp(-\mu t_I)$ (i.e., an exponentially distributed infectious period is assumed) and $R(\theta) = R_0 \mu t_I$ (i.e., the expected number of transmissions by an infected host is proportional to their infectious period). Substituting into Eq. (S4) then gives

$$p_{\text{outbreak}} = 1 - \int_0^{\infty} \mu \exp(-(1 + R_0 p_{\text{outbreak}}) \mu t_I) \, \mathrm{d}t_I.$$

Integrating and taking the largest solution between 0 and 1 of the resulting quadratic equation then reproduces the well-known formula,

$$p_{\text{outbreak}} = \max\left\{1 - \frac{1}{R_0}, 0\right\}.$$

Similarly, the outbreak risk under branching process approximations of a wide range of more complex compartmental models, for example models with non-exponentially distributed infectious periods and/or age structure, can also be represented using Eq. (S4).

### Supplementary Note 4: Outbreak risk under delayed and/or time-limited regular antigen testing

Here, we generalise our results to obtain an expression for the outbreak risk in scenarios where regular antigen testing is introduced reactively after an infection occurs and/or is only in place for a limited period of time. For simplicity, we consider homogeneous within-host dynamics (although the derivation presented here readily generalises to a heterogeneous population), supposing that each host infected at calendar time $t$ transmits the pathogen at rate $\beta(\tau, t)$ at time since infection $\tau$ (i.e., at calendar time $(t+\tau)$). Below, we first derive the outbreak risk for general $\beta(\tau, t)$, before deriving a specific form of $\beta(\tau, t)$ under delayed and/or time-limited regular antigen testing.

We suppose that an infected individual, who was infected at calendar time $t$, is introduced into an otherwise uninfected population at time since infection $\tau$ (at calendar time $(t+\tau)$). Then, conditioning on whether or not the initial infected host transmits the pathogen between times since infection $\tau$ and $(\tau + \mathrm{d}\tau)$ (and assuming no more external infections), we find that the extinction probability, $q(\tau, t)$, satisfies (up to terms of order $\mathrm{d}\tau^2$)

$$q(\tau,t) = q(\tau + \mathrm{d}\tau, t)q(0, t+\tau+\mathrm{d}\tau) \times \beta(\tau,t)\mathrm{d}\tau + q(\tau+\mathrm{d}\tau, t) \times (1-\beta(\tau,t)\mathrm{d}\tau).$$

Rearranging and taking the limit $\mathrm{d}\tau \to 0$ gives the differential equation,

$$\frac{\partial q}{\partial \tau} = q(\tau,t)\big(1 - q(0, t+\tau)\big)\beta(\tau,t),$$

which can be solved alongside the boundary condition $q(\infty, t) = 1$ to obtain

$$q(\tau,t) = \exp\left(-\int_{\tau}^{\infty}\big(1 - q(0, t+\tilde{\tau})\big)\beta(\tilde{\tau},t)\mathrm{d}\tilde{\tau}\right).$$

In particular, we have

$$q(0,t) = \exp\left(-\int_{0}^{\infty}\big(1 - q(0, t+\tau)\big)\beta(\tau,t)\mathrm{d}\tau\right),$$

(where we have relabelled $\tilde{\tau}$ from the previous equation as $\tau$) and the outbreak risk, $p_{\text{outbreak}}(t) = 1 - q(0,t)$, following the introduction of a single newly infected host at time $t$, is therefore the largest solution between 0 and 1 of the equation

$$p_{\text{outbreak}}(t) = 1 - \exp\left(-\int_{0}^{\infty} p_{\text{outbreak}}(t+\tau)\beta(\tau,t)\mathrm{d}\tau\right). \quad \text{(S5)}$$

We now derive the form of $\beta(\tau,t)$ under delayed and/or time-limited regular antigen testing. Specifically, we suppose that testing only takes place between calendar times $t_{\text{start}}$ and $t_{\text{end}}$, and that within that time period, the interval between tests (for a specified individual) is exponentially distributed with mean $T$. A similar argument to that in **Supplementary Note 1** can be used to show that the probability, $p_d(\tau,t)$, of an individual infected at calendar time $t$ having been detected by time since infection $\tau < \tau_{\text{inc}}$ (where $\tau_{\text{inc}}$ is the incubation period, with $p_d(\tau,t) = 1$ for $\tau \geq \tau_{\text{inc}}$), is

$$p_d(\tau,t) = 1 - \exp\left(-\frac{1}{T}\int_{\max\{0,\,\min\{\tau,\,t_{\text{start}}-t\}\}}^{\max\{0,\,\min\{\tau,\,t_{\text{end}}-t\}\}} p_+\big(V(x)\big)\mathrm{d}x\right),$$

where $p_+(V(x))$ gives the probability that the result of a test taken at time since infection $x$ is positive, and the limits of the integral give the times since infection up to $\tau$ over which the regular antigen testing policy is in place. The (calendar time-dependent) expected infectiousness profile, accounting for different possible detection times, is then

$$\beta(\tau,t) = \left[\alpha_d p_d(\tau,t) + \big(1 - p_d(\tau,t)\big)\right] \times \beta_u(\tau),$$

where $\beta_u(\tau)$ is the infectiousness profile of an undetected individual at time since infection $\tau$, and $\alpha_d$ is the relative infectiousness of a detected host.

Finally, we consider a scenario in which regular antigen testing is introduced after a delay of $x_{\text{del}}$ from the time of the first infection (where we may expect $x_{\text{del}}$ to be at least the length of the incubation period), and is carried out over a finite duration of time, $x_{\text{dur}}$. This scenario can be represented by taking $t_{\text{start}} = 0$ and $t_{\text{end}} = x_{\text{dur}}$ in the above, and then using Eq. (S5) to calculate $p_{\text{outbreak}}(-x_{\text{del}})$ numerically. In this scenario, for $t \geq t_{\text{end}}$, $p_{\text{outbreak}}(t)$ is simply the outbreak risk in the absence of antigen testing (which is independent of $t$ and can be calculated using Eq. (1) in the main text). Eq. (S5) can therefore be solved iteratively on a grid of $t \in [-x_{\text{del}}, t_{\text{end}}]$ by considering successively lower $t$ values and each time discretising the integral in Eq. (S5) to calculate $p_{\text{outbreak}}(t)$, since Eq. (S5) allows $p_{\text{outbreak}}(t)$ to be calculated once $p_{\text{outbreak}}(x)$ is known for $x > t$.

We note that the above numerical scheme in fact gives the outbreak risk for a grid of delays from 0 up to and including $x_{\text{del}}$ (for a fixed duration, $x_{\text{dur}}$). If there is no delay, then it is convenient to instead take $t_{\text{start}} = -\infty$ and $t_{\text{end}} = 0$, and to calculate $p_{\text{outbreak}}(t)$ on a grid of negative $t$ in order to obtain the outbreak risk for different durations of regular antigen testing (although we note that this scenario is unlikely to be of real-world relevance, unless an infection occurs shortly before the scheduled end of an ongoing regular antigen testing program).

### Supplementary Note 5: Details of discrete-time stochastic outbreak simulation algorithm

We verified our analytically derived estimates of the outbreak risk by comparing these estimates with corresponding estimates obtained through repeated simulation of a discrete-time, individual-based stochastic epidemic model (**Figure 2F** and **Supplementary Figure 2**). In this section, we describe the simulation model and how it was used to estimate the outbreak risk.

Prior to running each outbreak simulation, we first determined and discretised the within-host dynamics that each individual, $i$, in the population would follow if ever infected, according to the following steps:

1. Determine the individual's (continuous-time) viral load profile, $V^{(i)}(\tau)$, where $\tau$ is the time since infection, and their incubation period, $\tau_{\mathrm{inc}}^{(i)}$ (in **Figure 2F** and **Supplementary Figure 2,** we assumed homogeneous within-host dynamics, but in principle heterogeneity could be included).
2. Calculate the individual undetected infectiousness profile, $\beta_u^{(i)}(\tau)$, and probability of a test taken at time since infection $\tau$ giving a positive result, $p_+^{(i)}(\tau)$, as described in the main text (note that $p_+^{(i)}$ is here defined as a function of time since infection rather than viral load).
3. Sample the (potential) time, $r^{(i)}$, from the start of the day of infection to the exact (potential) infection time, uniformly between zero and one day.
4. For each day since infection, $\tau_{\mathrm{discr}} \geq 1$ (where the day of infection is denoted day 0), calculate the discretised infectiousness, $\beta_{u,\mathrm{discr}}^{(i)}(\tau_{\mathrm{discr}})$, as the average value of $\beta_u^{(i)}(\tau)$ between times since infection $(\tau_{\mathrm{discr}} - r_i)$ and $(\tau_{\mathrm{discr}} - r_i + 1)$. Note that implicit in our simulation algorithm is the assumption that hosts cannot transmit the pathogen on the day of infection (i.e. $\beta_{u,discr}^{(i)}(0) = 0$).
5. Calculate the probability, $p_{+,\mathrm{discr}}^{(i)}(\tau_{\mathrm{discr}}) = p_+^{(i)}(\tau_{\mathrm{discr}} - r_i)$, of a test taken at the start of day of infection $\tau_{\mathrm{discr}} \geq 1$ giving a positive result.
6. Calculate the discretised incubation period, $\tau_{\mathrm{inc,discr}}^{(i)} = \left\lfloor \left( r^{(i)} + \tau_{\mathrm{inc}}^{(i)} \right) \right\rfloor$. Note that we only considered a single continuous incubation period, which exceeded 1, but if a non-trivial distribution is used, then it should be truncated to take values of at least 1 in order to avoid symptom onset occurring on the day of infection.

7. Calculate the relative infectiousness on the day of symptom onset (assuming the host is not detected before developing symptoms), $\alpha_o^{(i)}$, chosen to ensure that the continuous- and discrete-time infectiousness profiles give the same expected number of transmissions during this day (under the assumption of isolation immediately following the exact symptom onset time).
8. Calculate the total duration of infection (up to loss of infectiousness), $\tau_{\mathrm{rec,discr}}^{(i)}$, as the earliest day of infection for which $\beta_u^{(i)}(\tau_{\mathrm{discr}}) = 0$ for all $\tau_{\mathrm{discr}} \geq \tau_{\mathrm{rec,discr}}^{(i)}$.

An example discretised infectiousness profile (without regular antigen testing) is shown in **Supplementary Figure 2A**.

In the simulation algorithm, individuals are classified as being in one of the following states on each day: susceptible ($S$), infected but undetected ($U$), infected with symptom onset on the current day (and not detected prior to onset; $O$), infected and detected ($D$), or recovered (specifically, no longer infectious following an infection; $R$). The $O$ stage is included to allow for symptom onset (and therefore detection) occurring at any time of day, whereas for simplicity we assumed that regular antigen testing takes place only at the start of each day. The status of individual $i$ (at a given step in the simulation) is denoted by $Y^{(i)} \in \{S, U, O, D, R\}$. We write, for example, $\mathbf{1}_S\left(Y^{(i)}\right)$, to denote the indicator function that takes the value 1 if $Y^{(i)} = S$, and 0 otherwise. However, we emphasise that the simulation model is not a compartmental model, since different individuals of the same status are not treated identically.

Now, the simulation algorithm has the following inputs:

- The population size, $N$.
- The relative infectiousness of detected hosts, $\alpha_d$.
- The relative susceptibility, $\eta^{(i)}$, of each individual, $i$ (note that we assumed homogeneous susceptibility in our analyses, i.e., $\eta^{(i)} = 1$ for each $i$).
- The quantities $\beta_{\mathrm{u,discr}}^{(i)}(\tau_{\mathrm{discr}})$, $p_{+,\mathrm{discr}}^{(i)}(\tau_{\mathrm{discr}})$, $\tau_{\mathrm{inc,discr}}^{(i)}$, $\alpha_o^{(i)}$ and $\tau_{\mathrm{rec,discr}}^{(i)}$, which characterise discretised individual within-host dynamics (as described above).
- The number of antigen tests, $Z^{(i)}(t)$, conducted by individual $i$ at the start of day $t$ of the simulation (for each positive integer value of $t$). We considered two possibilities:

i. In **Figure 2F**, we sampled $Z^{(i)}(t)$ from a Poisson distribution with mean $1/T$ (independently for each individual and each day, where a range of $T$ values were considered). This is consistent with our analytic outbreak risk derivation (since an exponentially distributed duration between tests, with mean $T$ (measured in days), leads to a Poisson-distributed number of tests being taken each day, with mean $1/T$, although note that tests may be taken at any time of day in the analytic approach), but leads to the possibility of more than one daily test.

ii. In **Supplementary Figure 3B**, we instead considered a fixed (constant) gap of length $T$ between days on which a test is taken. For each individual, we sampled the first day of the simulation on which a test is conducted uniformly between 1 and $T$ (independently for each individual).

The outbreak simulation algorithm consists of the following steps:

1. Initialise the time at $t = 0$ days and the status of each host at $Y^{(i)} = S$.
2. Sample a single initial infected host, $i$, according to the relative susceptibilities, $\eta_j$ (i.e., host $j$ is selected with probability $\eta_j / \sum_{k=1}^{N} \eta_k$). Set $Y^{(i)} = U$ and the infection time, $t_{\text{inf}}^{(i)} = 0$.
3. While $\sum_{i=1}^{n} \mathbf{1}_{\{U,O,D\}}\left(Y^{(i)}\right) > 0$ (i.e., while the number of active infections is greater than zero), repeat the following steps:
   a. Increase the simulation time, $t$, by 1 day (i.e., set $t = (t+1)$).
   b. For each $i$ such that both $Y^{(i)} \in \{U, O, D\}$ and $\left(t_{\text{inf}}^{(i)} + \tau_{\text{rec,discr}}^{(i)}\right) = t$, set $Y^{(i)} = R$ (recovery/loss of infectiousness).
   c. For each $i$ such that $Y^{(i)} = O$, set $Y^{(i)} = D$ (day after symptom onset).
   d. For each $i$ such that $Y^{(i)} = U$, carry out the following steps (testing process):
      i. Generate a random number, $r$, uniformly distributed between 0 and 1.
      ii. If $r < 1 - \left(1 - p_{+,\text{discr}}^{(i)}\left(t - t_{inf}^{(i)}\right)\right)^{Z^{(i)}(t)}$, set $Y^{(i)} = D$.
   e. For each $i$ such that both $Y^{(i)} = U$ and $\left(t_{\text{inf}}^{(i)} + \tau_{\text{inc,discr}}^{(i)}\right) = t$, set $Y^{(i)} = O$ (symptom onset).
   f. Calculate the total infectious pressure exerted on each susceptible individual over the current simulation day,

$$\lambda(t) = \frac{1}{N}\left(\sum_{j=1}^{N}\left(\mathbf{1}_U\left(Y^{(j)}\right) + \alpha_o^{(j)}\mathbf{1}_O\left(Y^{(j)}\right) + \alpha_d\mathbf{1}_D\left(Y^{(j)}\right)\right)\beta_{\text{u,discr}}^{(j)}(t - t_{inf}^{(j)})\right).$$

g. For each $i$ such that $Y^{(i)} = S$, carry out the following steps (transmission process):

i. Generate a random number, $r$, uniformly distributed between 0 and 1.

ii. If $r < 1 - \exp(-\eta^{(i)}\lambda(t))$, set $Y^{(i)} = U$ and $t_{inf}^{(i)} = t$.

For each testing scenario considered, we carried out 100,000 model simulations in a population of $N = 1{,}000$ individuals. The outbreak risk was estimated as the proportion of model simulations in which the total number of individuals ever infected exceeded 10% of the total population (see **Supplementary Figure 2**).

### Supplementary Table 1

| Parameters | Symbol | Unit | Value | How obtained |
|---|---|---|---|---|
| Rate constant for virus infection | $b$ | (copies/ml) $^{-1}$ day$^{-1}$ | $1.43 \times 10^{-7}$ | Fitted to viral load data |
| Maximum rate constant for viral replication | $\gamma$ | day$^{-1}$ | 5.64 | Fitted to viral load data |
| Death rate of infected cells | $\delta$ | day$^{-1}$ | 1.21 | Fitted to viral load data |
| Incubation period | $\tau_{\mathrm{inc}}$ | days | 4.60 | Fitted to viral load data |
| Initial quantity of free virus | $V(0)$ | copies/ml | 0.01 | Assumed (3) |
| Standard deviation of error in log viral load measurements | σ | $\log_{10}$ (copies/ml) | 0.87 | Fitted to viral load data |
| Limit of infectiousness and detection limit of antigen test | $V^*$ | $\log_{10}$ (copies/ml) | 3.30 | Minimum viral load for culturable virus for the omicron variant from (4) |
| Relative infectiousness of detected hosts | $\alpha_{\mathrm{d}}$ | --- | 0.26 | Estimated value for the delta variant from (5) (other values considered in **Figure 3**) |
| Reproduction number at time of introduction in absence of regular antigen testing | $R_0$ | --- | 1.5 | Assumed (other values considered in **Figure 3** and elsewhere) |
| Mean interval between antigen tests when regular testing conducted | $T$ | days | 2 | Assumed (**Figure 2BC** only; a range of values considered elsewhere) |

**Supplementary Table 1. Baseline parameter values used in our analyses.** These parameter values were used in our analyses except where explicitly stated otherwise. Note that the values of the within-host model parameters $b$, $\gamma$, $\delta$ and $\tau_{inc}$ here are population median estimates (fixed effects); estimates of random effects are given in **Supplementary Table 2** (the random effects were used to account for heterogeneous within-host dynamics in **Figure 4**).

### Supplementary Table 2

| Parameters | Symbol | Value |
|---|---|---|
| Random effect for rate constant for virus infection | $\omega_b$ | 1.33 |
| Random effect for maximum rate constant for viral replication | $\omega_\gamma$ | 0.15 |
| Random effect for death rate of infected cells | $\omega_\delta$ | 0.54 |
| Random effect for incubation period | $\omega_{\tau_{\mathrm{inc}}}$ | 0.29 |

**Supplementary Table 2. Estimated random effects for within-host model parameters.** The estimated quantities correspond to the standard deviation of the natural logarithm of the parameters $b$, $\gamma$, $\delta$ and $\tau_{inc}$ between different individuals (see the section "Within-host model and parameter estimation" of **Methods** in the main text for details; population parameter values (fixed effects) and units are given in **Supplementary Table 1**).

### Supplementary Figure 1

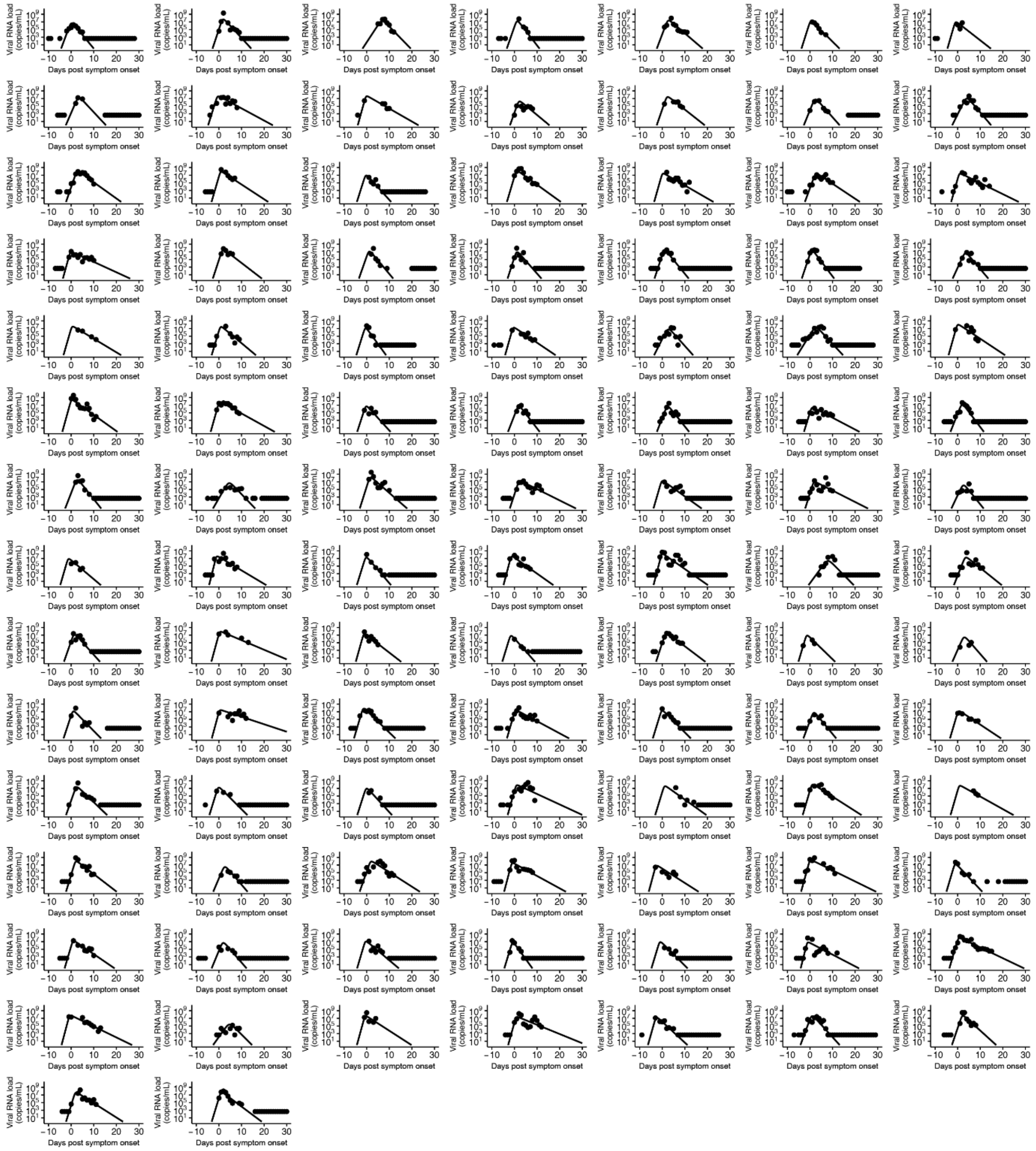


**Supplementary Figure 1. Reconstructed viral dynamics for individual hosts.** Individual-level model fits to longitudinal SARS-CoV-2 viral load data using a target cell-limited within-host model are shown (see the section “Within-host model and parameter estimation” of **Methods** in the main text). Overall, we used data from 521 individuals with omicron variant infections (6) to characterise SARS-CoV-2 viral dynamics; here, individual model fits are shown for 100 randomly chosen individuals. In each panel (corresponding to a single individual), the dots indicate the measured viral load data, and the solid curves the estimated viral load at different times relative to symptom onset.

### Supplementary Figure 2

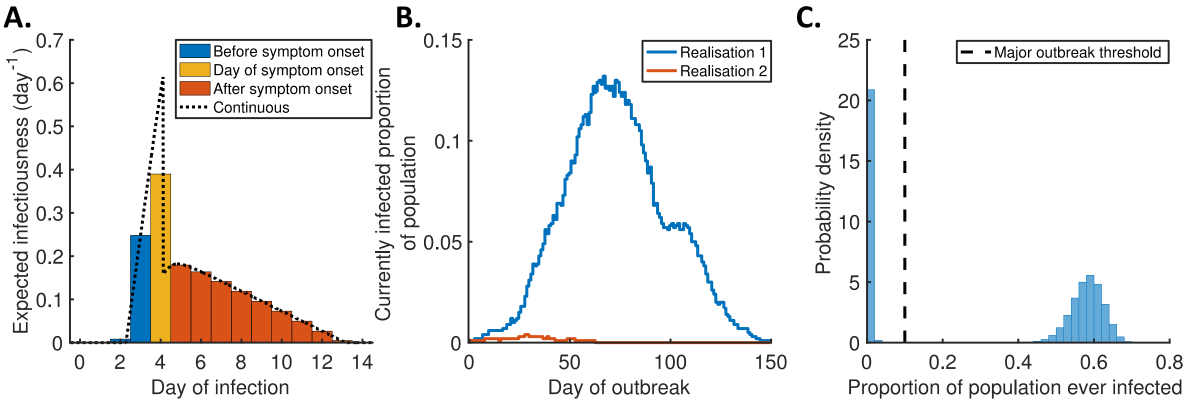


**Supplementary Figure 2. Alternative estimation of the outbreak risk using a discrete-time, individual-based, stochastic outbreak simulation model. A**. Example discretised infectiousness profile of a single infected host when regular antigen testing does not take place. **B**. The output of two realisations of the stochastic outbreak simulation model. **C**. Histogram of total outbreak sizes (i.e., the total proportion of the population ever infected during the outbreak) over 100,000 model simulations. The vertical black dashed line indicates the assumed threshold for a major outbreak of 10% of the population being infected. Here (without regular antigen testing), the estimated outbreak risk (i.e., the proportion of model simulations classified as major outbreaks) is 0.58. Details of the stochastic simulation model are given in **Supplementary Note 5**.

**Supplementary Figure 3**

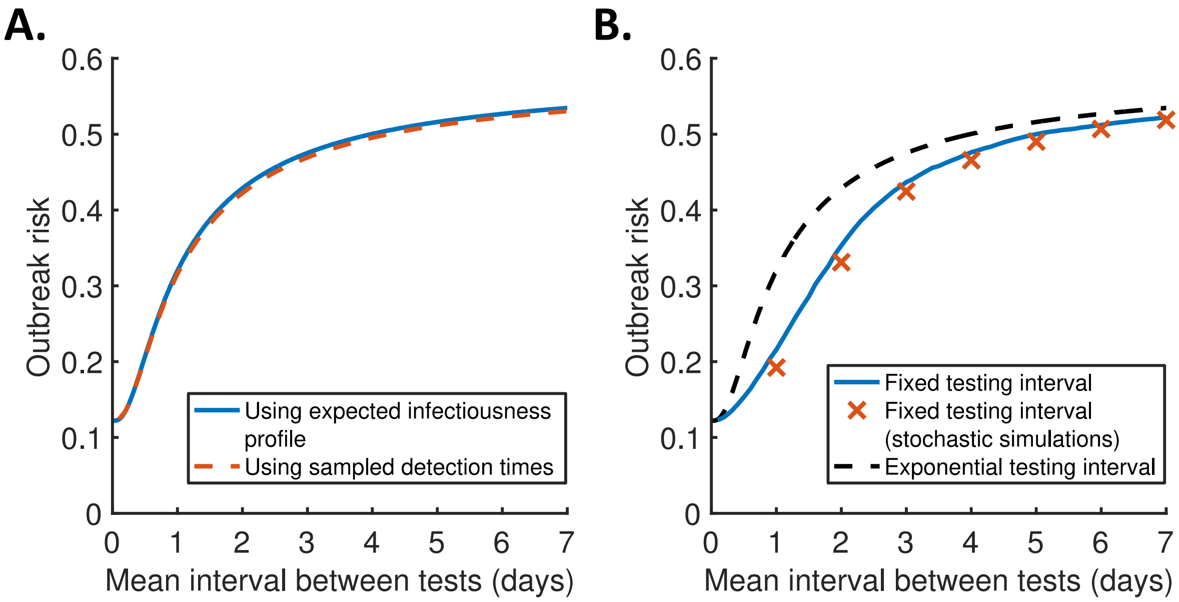


**Supplementary Figure 3. Effect of details of implementation of antigen testing in our modelling approach on the outbreak risk under regular antigen testing. A.** The outbreak risk for different values of the (mean) interval between antigen tests, comparing our default analytic approach using an expected infectiousness profile that averages over the individual infectiousness profiles of hosts with different detection times (blue), and a more complex approach in which variations in detection times are accounted for directly by sampling the detection times of a large number of individuals and using Eq. (2) to calculate the outbreak risk (red dashed). **B.** The outbreak risk for different values of the (mean) interval between tests, comparing our default analytic approach assuming an exponentially distributed interval between tests (black dashed), and both the analytic (blue) and simulation-based approaches (red crosses) under the alternative assumption of a fixed (constant) interval between tests. Note that in the analytic approach with a fixed interval, we used sampled detection times since the expected infectiousness profile was not readily available in this case.

### Supplementary Figure 4

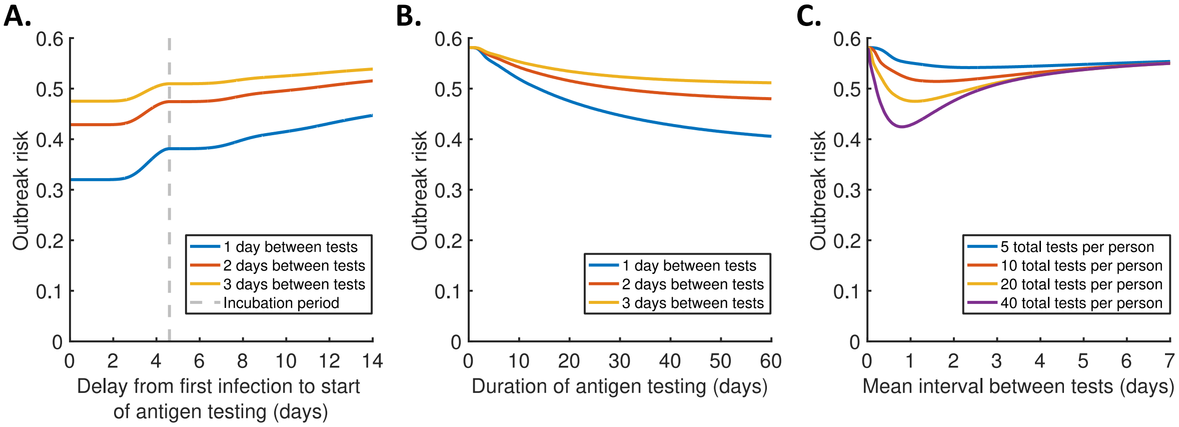


**Supplementary Figure 4. Effect of delayed and/or time-limited antigen testing. A**. The outbreak risk for different delays from the time of the first infection to the introduction of regular antigen testing, assuming an infinite duration of testing, and either 1 (blue), 2 (red) or 3 days (orange) between tests (on average). **B**. The outbreak risk for different durations of antigen testing, assuming a delay of one incubation period (4.6 days) from the first infection to the start of testing (i.e., testing starts following the detection of a symptomatic case), and either 1 (blue), 2 (red) or 3 days (orange) between tests (on average). **C**. The outbreak risk for different values of the (mean) interval between tests with a fixed total of 5 (blue), 10 (red), 20 (orange) or 40 (purple) tests available to each individual (on average), assuming a delay of one incubation period from the first infection to the start of testing.
